# Physicians’ Attitudes towards Secondary Use of Clinical Data for Biomedical Research Purposes in Germany. Results of a Quantitative Survey

**DOI:** 10.1101/2022.08.22.22279095

**Authors:** Anja Köngeter, Christoph Schickhardt, Martin Jungkunz, Katja Mehlis, Eva C. Winkler

## Abstract

**Background:** For biomedical data-driven research purposes, secondary use of clinical data carries great but largely untapped potential. Physicians’ attitudes and their needs towards secondary data use are essential to inform its practical and ethically sound implementation but are currently understudied.

**Objective:** Therefore, the objectives of the study are to assess physicians’ (i) general attitudes and concerns, (ii) willingness to adapt workflows and to make data available for secondary use, (iii) group-specific conditions and concerns of physician-scientists and purely clinical physicians.

**Methods:** We developed an online survey based on a literature review and an expert interview study. Physicians in private practice and at two large German university hospitals were surveyed from May 2021 until January 2022.

**Results:** In total, 446 physicians participated in the survey. 96% [380/397] of all physicians reported a positive attitude towards secondary use; 87% [31/397] are in-principle willing to support secondary use of clinical data along with a small proportion of physicians with fundamental reservations 8%. Secondly, the most important conditions for adapting workflows was funding of additional time and effort for research-adequate documentation (71% [286/390]) and the most important condition for providing patients’ clinical data was reliable protection of patients’ privacy (67% [254/382]). Thirdly, physician-scientists were more likely to request additional funding for research-adequate documentation as a precondition for support (83% vs 69%, *P*=.002) and the privilege to conduct research with patient data prior to other researchers (43% vs 11%, *P*<.001); while purely clinical physicians more frequently require reliable protection of patient privacy (76% vs 62%, P=.007) and monetary compensation (45% vs 25%, *P*<.001).

**Conclusion:** Since this study presents high in-principle willingness of physicians to support secondary use along with little general concerns, it seems essential to address physicians’ group-specific conditions toward secondary use in order to gain their support.

## Introduction

Secondary use of clinical data for biomedical research purposes holds promising potential for various types of non-interventional, data-driven research. We define secondary use as the collection and reuse of clinical data in data-gathering, non-interventional biomedical research and quality improvement activities (1). Research with clinical data collected in routine care bears the advantage of not involving additional physical interventions or the collection of additional data. In Germany, the medical informatics initiative, funded by the German Federal Ministry of Education and Research, aims at fostering secondary use of clinical data from health care facilities (2). Although secondary use aims to improve biomedical knowledge and thus medical care, it is highly unlikely to directly benefit the patients who released their data.

Physicians’ attitudes towards secondary use have hardly been investigated. Some studies have focused on related topics. For example, a systematic literature review about researchers’ and health care professionals’ perspectives on data sharing of clinical trial data and health administrative data pinpoints major concerns related to privacy, access to data, and potential for misinterpretation of data (3); data to be shared are defined differently in these studies, and sometimes are not clearly characterized. Several *qualitative* studies highlight concerns related to secondary use of different kinds of health data in general practitioners (GPs) (4-7) and oncologists (8). So far, it is unclear how pronounced and widespread these concerns are.

The scarce socio-empirical evidence is unsatisfying given the relevance of physicians’ support for the implementation of new workflows as suggested by an exploratory interview study we conducted in preparation for the present survey: experts considered willingness of physicians to provide support for the implementation of secondary use in German hospitals and private practices both critical and possibly insufficient (9); one GP stated that peers would be reluctant to change their work routine. Regarding monetary incentives to change work routine, two qualitative studies of Australian GPs come to contradictory conclusions (4, 5). To our knowledge, there are no quantitative studies that examine whether and under what conditions physicians are willing to adjust their work routine for secondary use.

So far, group distinctions between physician-scientists and purely clinical physicians have not been examined. However, two studies indicate disparities between these groups: While Canadian health researchers widely accepted secondary use for research (10) whereas Canadian GPs showed a far lower approval rate (11).

The aim of this study is therefore to assess physicians’ attitudes and group-specific needs regarding secondary use which can inform its practical implementation. To address the delineated research gap, the objectives of the present study are to assess physicians’ (i) general attitudes and concerns, (ii) in-principle willingness to adapt workflows to share data for secondary use, (iii) group-specific conditions to support secondary use in physician-scientists and purely clinical physicians. To our knowledge, we present the first quantitative analysis of physicians’ conditions to support secondary use of clinical data for research purposes and the first study differentiating between physician-scientists’ and purely clinical physicians’ needs.

## Methods

### Survey Development

The questionnaire is based on a literature review and the results of an expert interview study (9). The expert interviews indicated distinct perceptions and expectations of physician-scientists and purely clinical physicians. Therefore, we derived group-specific hypotheses that we operationalized in the form of questionnaire items. The questionnaire was developed and discussed with members of an interdisciplinary research team consisting of social scientists, ethicists, legal scholars, and physicians. To ensure comprehensibility and technical function of the 22 item questionnaire, we pretested the survey by cognitive interviews (n=6) with physicians with and without experience in generating and using clinical data for research purposes. Based on the results, we adjusted the wording of the specific conditions for physicians’ support to improve comprehensibility.

The survey included information about risks and benefits to patients associated with secondary use as well as the potential increase in documentary workload for clinicians to allow participants to develop an informed opinion. The self-administered, anonymous, online survey covered attitudinal questions designed as 5-point Likert scale. Measurement of pronounced research interest was operationalized by asking participants define themselves as physician-scientists; participants who do not define themselves as physician-scientist were referred to as purely clinical physicians. The study obtained ethics approval from the University of Heidelberg’s research ethics committee (reference number S-361/2018). This survey was approved by the data protection officer of Heidelberg University Hospital. These approvals were valid for all data collections.

### Sampling and recruitment

The survey was administered via three data collections: For the first data collection the Cancer Registry of the German federal state of Baden Wuerttemberg sent e-mail invitations to physicians in Baden Wuerttemberg who had reported more than two patients in the registry until April 2021 (census, N=3.313). The 2^nd^ and the 3^rd^ data collections complemented the first and was targeted to physicians at university hospitals and thereby a research-oriented environment. For the *2*^*nd*^ *data collection* at the Heidelberg University Hospital (census, N=1,686) an e-mail distribution list of all physicians with patient contact was created and was authorized by the university hospital board and the employee council. And for the *3rd data collection* all physicians of the Charité - University Hospital Berlin were invited via email by the Charité-BIH Clinical Study Center (census, N=3,870). All e-mail invitations contained a weblink to the anonymous survey.

Individuals who completed the survey were not compensated. Data collections occurred from May 2021 until January 2022 with a duration of 3-4 weeks each. For the 2^nd^ and 3^rd^ data collection, an e-mail reminder was sent out 8 days after the first invitation.

### Analysis

Descriptive statistics were used to express categorical variables as counts and percentages. Differences in proportions were assessed for statistical significance (*P<* 0.05) by way of χ2 tests. Significances of group differences in mean values were calculated using the Mann-Whitney U test (two-tailed). All analyses were performed using SPSS IBM version 28.

## Results

### Participant characteristics

Of the 8,615 physicians contacted, 446 responded to the survey (response rate: 5%); after excluding participants who answered less than 50% of the items or dropped out before item no. 11, the dataset used for analysis encompassed 397 cases. Of these included physicians, 79% [313/397] worked at a university hospital and 15% [60/397] in a private practice (Table 1). Gender distribution was balanced. 62% [245/397] reported more than 10 years of work experience’.

**Table 1.** Demographics of participants (n=397)

| Characteristics | Values, n (%) |
| --- | --- |
| **Affiliation** |  |
| University Hospital / Academic Teaching Hospital | 313 (78.64) |
| Private practice | 60 (15.08) |
| Public community hospitals and for-profit hospitals | 20 (5.03) |
| n/a | 4 (1.26) |
| **Years of Service** |  |
| ≤ 10 | 150 (37.69) |
| > 10 | 245 (61.56) |
| n/a | 2 (0.50) |
| **Medical Speciality (top five)** |  |
| Internal Medicine | 88 (22.11) |
| Surgery | 39 (9.80) |
| Gynecology | 39 (9.80) |
| Anesthesiology | 29 (7.29) |
| Pediatrics and Youth Medicine. | 25 (5.53) |
| **Sex** |  |
| Female | 180 (45.23) |
| Male | 207 (52.01) |
| n/a | 10 (2.52) |
| **Identifies as physician-scientist** |  |
| Yes | 251 (63.07) |
| No | 125 (31.41) |
| n/a | 21 (5.29) |
| **Involved in conducting studies using clinical data from patient care within the last 5 years** |  |
| Yes | 321 (80.65) |
| No | 72 (18.09) |
| n/a | 4 (1.01) |

Overall, 63% [251/397] indicated perceiving themselves as physician-scientist; at university hospitals 84% [243/294] of all physicians considered themselves physician-scientists. 31% [125/397] were defined as purely clinical physicians by indicating that they do not perceive themselves as physician-scientists; 98% [57/58] of all physicians working in private practice were purely clinical physicians. Of all participants, 81% [321/397] reported having contributed to studies using clinical data in the last 5 years.

### General attitudes towards supporting secondary use

With 96% [380/397] almost all respondents deemed secondary data use for research purposes important and 68% [269/397] hold the view that, as a physician, they have a moral obligation to provide clinical data from patient care for research purposes (Fig 1). Only 8% [31/397] of the participants had fundamental reservations. Yet, 13% [50/397] of respondents were concerned that patients would report fewer details about their illness and 11% [41/397] of physicians would document differently to protect their patients’ sensitive information (e.g. stigmatizing data) against misuse; purely clinical physicians expressed significantly stronger fundamental reservation (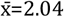 vs 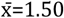 *P*<.001), were more concerned that their patients will report fewer details (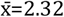 vs 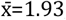, *P*=.001), and were more inclined to change their way of documentation (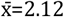 vs 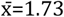, *P*=.001) compared to physician-scientists *(Table 2)*.

**Table 2:** Significance of differences in general attitudes toward secondary use and willingness to support secondary use by self-perception as physician-scientist and purely clinical physician.

| Agreement to Statement | Physician scientist, mean (SD) | Purely clinical physician, mean (SD) | P value<sup>a</sup> |
| --- | --- | --- | --- |
| I think it is fundamentally important that patients' clinical data be used for research purposes. (n = 397) | 4.90 (0.41) | 4.53 (0.85) | <.001 |
| I am generally willing to support research using my own patients' clinical data. (n = 397) | 4.66 (0.81) | 4.12 (1.14) | <.001 |
| As a physician, I have a moral obligation to provide clinical data from patient care for research purposes (if patients consent to this use). (n = 397) | 4.12 (1.12) | 3.56 (1.27) | <.001 |
| In principle, I am prepared to implement any necessary adjustments to workflows in documentation for research purposes as part of my job. (n = 390) | 4.14 (1.00) | 3.56 (1.16) | <.001 |
| In principle, I am prepared to implement any necessary adjustments to workflows with regard to information and consent processes as part of my job. (n = 390) | 4.09 (1.02) | 3.48 (1.19) | <.001 |
| I have fundamental reservations about supporting research with clinical data from my own patients. (n = 397) | 1.50 (0.91) | 2.04 (1.22) | <.001 |
| I am concerned that my patients will report fewer details about their conditions if they know that their clinical data will be used for research purposes. (n = 397) | 1.93 (1.05) | 2.32 (1.16) | .001 |
| If my patients' clinical data is collected for research, I will document differently to protect my patients' data against misuse (e.g., I will not document stigmatizing diagnoses). (n = 397) | 1.73 (1.03) | 2.12 (1.21) | .001 |

<sup>a</sup> Significances of group differences in mean values were calculated using the Mann-Whitney U test; Values in italics are significant at .05 level of significance (two-tailed); 5 Point Likert Scale: Disagree 1; Agree 5.

**Figure 1:**
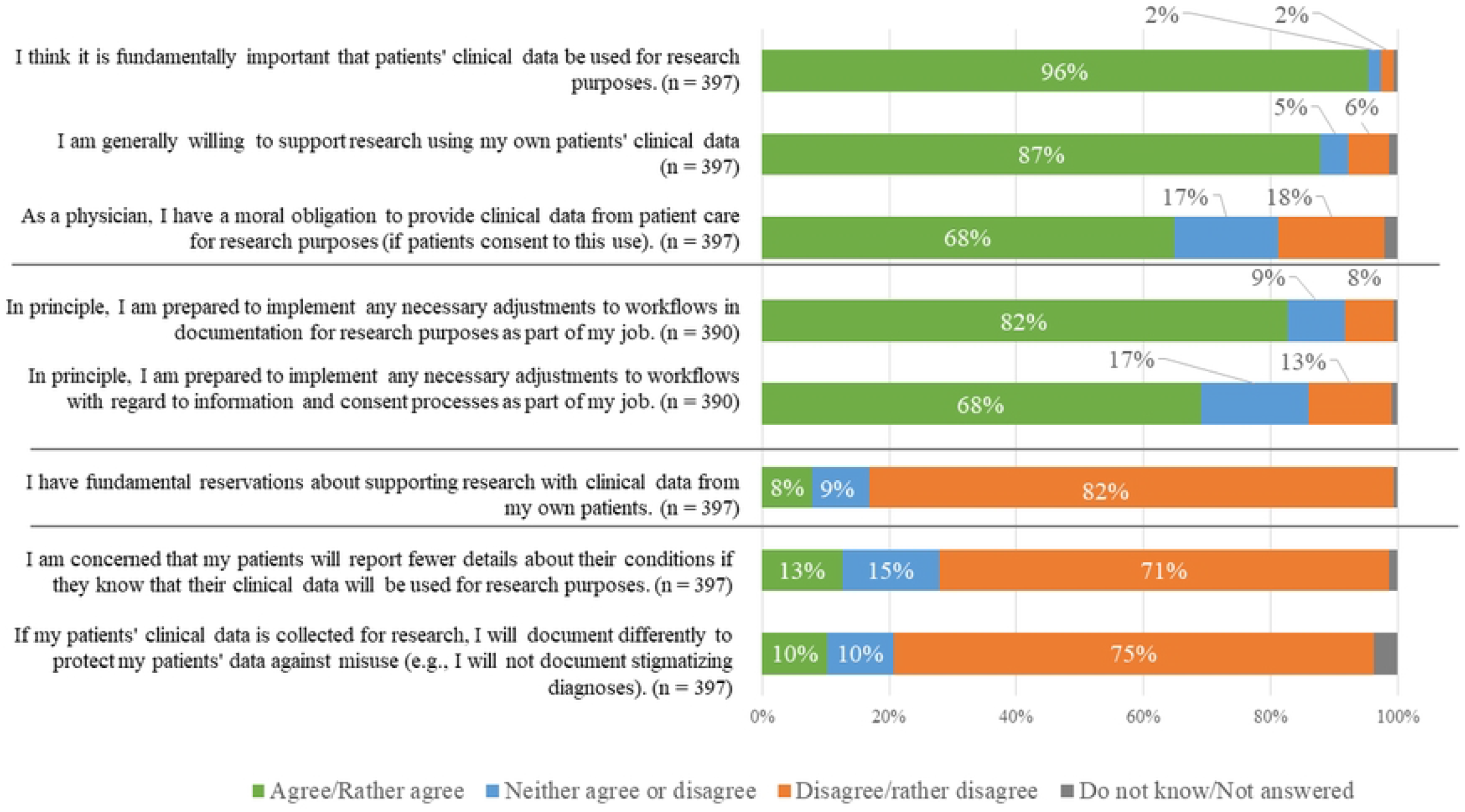
General attitudes and concerns of physicians towards secondary use of clinical data for research purposes and their willingness to support secondary use.

87% [348/390] of all surveyed physicians were willing to support secondary research use in principle. When being asked about potentially necessary adjustments of the workflow, 71% [286/390] were willing to implement changes for documentation, and 67% [269/390] were willing to inform and consent patients for secondary use. In contrast to purely clinical physicians, physician-scientists were more convinced about the importance of secondary use (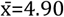 vs 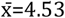, *P*<.001), more willing to adjust documentation (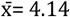 vs 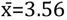, *P*<.001), and more willing to obtain informed consent (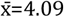 vs 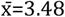, *P<*.001).

### Conditions for adapting workflows

Participants were then asked to indicate what seems most important to them for adapting their workflows in order to support secondary data use. They did so by selecting the three most important conditions out of a list. The most frequently selected conditions were funding of additional person-hours for research-appropriate documentation (77% [302/390]), funding of newly developed and user-friendly software to support research-appropriate documentation (62% [243/390]), and funding of additional person-hours for obtaining informed consent (60% [236/390]) *(Table 3)*. Physicians-scientists and purely clinical physicians differed in only one aspect; funding of additional person-hours for research-adequate documentation was more important to physician-scientists (83% vs 69%, *P*=.002).

**Table 3:** Conditions for adapting workflows for secondary use (documentation and obtain consent); significance of group differences (n=390)

|  | All participants, n (%) | Physician-scientists, n (%)<sup>b</sup> | Purely clinical physicians, n (%)<sup>b</sup> | *P value*<sup>c</sup> |
| --- | --- | --- | --- | --- |
| **Conditions to implement adaptations of the workflows<sup>a</sup>** |  |  |  |  |
| Funding for additional person-hours for research-appropriate documentation | 302 (77.44) | 205 (82.66) | 83 (68.60) | *.002* |
| Funding of newly developed, user-friendly software to support research-appropriate documentation | 243 (62.31) | 162 (65.32) | 67 (55.37) | .064 |
| Funding for additional person-hours for patient information related to consent | 236 (60.51) | 153 (61.69) | 71 (58.67) | .578 |
| Improving the quality of care across the healthcare system | 123 (31.54) | 77 (31.04) | 38 (31.40) | .945 |
| Improving the quality of care in one's own practice/department through feedback systems fed by the analysis of clinical data from all integrated practices/departments - including one's own practice/department | 89 (22.82) | 50 (20.16) | 35 (28.92) | .061 |
| Free access to publications based on routine care clinical data | 69 (17.69) | 40 (16.12) | 26 (21.48) | .207 |
| Certificate in recognition of generating high-quality clinical data for research purposes | 23 (5.90) | 17 (6.85) | 6 (4.95) | .479 |

<sup>a</sup> Multiple Answer item; maximum 3 answers.
<sup>b</sup> Of all participants who answered this item (n=390), n= 369 indicated whether or not they consider themselves physician-scientists (n=248) or physicians (n=121).
<sup>c</sup> Chi-Squared Test (Pearson); values in italics are significant at .05 level of significance.

When asked about participants’ willingness to spend additional time for research-adequate documentation per patient visit, 53% [165/314] of the participants was willing to document up to additional 2-6 minutes, 25% [79/314] of the participants to document up to 2 minutes, and 22% [70/314] was willing to document >=6 minutes. A significantly larger proportion of physician-scientists were willing to document >=6 minutes compared to purely clinical physicians (28% vs 11%, *P*=.005).

63% [249/386] of respondents favoured trained, non-medical staff to inform and consent patients about secondary use; 31% [123/386] felt themselves, as physicians, responsible for this task.

### Conditions for providing patients’ clinical data

All participants were asked about conditions they deem most important in order to make their patients’ clinical data available for secondary use. The most frequently reported conditions were: reliable protection of patients’ privacy (66% [254/382]), extra protection for sensitive data (e.g., genetic data, psychiatric data) (47% [179/382]), and notification about additional/incidental findings relevant to their patients’ health (37% [140/382]) (Table 3).

Patient privacy was significantly more often important to purely clinical physicians than to physician-scientists (76% vs 62%, *P*=.007) as was the monetary compensation for making research-adequate clinical data available (45% vs 25%, *P*<.001). For Physician-scientists the right to first conduct their own research with their patients’ clinical data was more relevant (43% vs 11%, *P*<.001) as was co-authorship in scientific articles based on clinical data of their patients (31% vs 8%, *P*<.001).

Being asked about the acceptance of potential data-users, 40% [159/381] of respondents agreed that all researchers, regardless of their affiliation, should be allowed to use their patients’ clinical data. 46% [174/381] were opposed to making clinical data available for researchers working for companies conducting medical research. 18% [67/381] did not want to provide data for collaborative projects between public research institutions and private companies.

Participants were asked about data ownership. They were divided on whether data can be owned (51%) or not (49%). Among those who held that data can be owned, 54% considered patients as data owners, and 40% believed that the practices and hospitals where data are collected or the physicians own the data, and only 6% thought the data belong to everyone who can use the data to add value to medical care. Purely clinical physicians were significantly more likely to think that data belong to patients (66% vs 48%, P=.024). Interestingly, the view that data can be owned was positively associated (eta= .191 P<.001) with the willingness to provide patients’ clinical data.

#### Concerns about provision of patient data

Participants were asked about their specific concerns with respect to the provision of their patients’ clinical data for secondary use. These concerns differed significantly between the two groups (*Table 4*). Compared to physician-scientists, purely clinical physicians were significantly more likely to have concerns about misuse of their patients’ clinical data through unauthorized access to datasets by third parties (75% vs 53%, *P*<.001), failure to protect their patients’ privacy (62% vs 36%, *P*<.001), and discrimination based on clinical data against their patients (27% vs 15%, *P*=.009). Purely clinical physicians were also more likely to have concerns about increased liability risk (e.g., research uncovers a misdiagnosis in their own practice/department) (36% vs 26%, *P*=.044), and a loss of patients’ trust in the physician-patient relationship (21% vs 12%, *P*=.019). The second most common concern of physician-scientists (after concerns about privacy of their patients) was insufficient data quality that can result in inaccurate findings (62% vs 42%, *P*<.001), and that other researchers conduct research using their patients’ clinical data before they do (43% vs 13%, *P*<.001).

**Table 4:** Conditions for providing patients’ clinical data available for medical studies and significance of differences in proportions by self-perception as physician-scientist or physician (n=382)

| Conditions to make your patients' clinical data available<sup>a</sup> | All participants, n (%) | Physician-scientists, n (%)<sup>b</sup> | Purely clinical physicians, n (%)<sup>b</sup> | P value<sup>c</sup> |
| --- | --- | --- | --- | --- |
| Reliable protection of the privacy of my patients | 254 (66.49) | 149 (61.57) | 91 (75.83) | .007 |
| Special protection measures for sensitive data (e.g. genetic data, psychiatric data) | 179 (46.85) | 108 (44.62) | 61 (50.83) | .265 |
| Notification of additional diagnostic findings in my patients that will be identified in the future as a result of planned research use of clinical data (for example, through novel methods of analysis of image data) | 140 (36.64) | 82 (33.88) | 50 (41.66) | .148 |
| Right to initially conduct my own research with my patients' clinical data | 125 (32.72) | 104 (42.97) | 13 (10.83) | <.001 |
| Monetary compensation for providing appropriately generated datasets suitable for research purposes | 122 (31.93) | 60 (24.79) | 54 (45.00) | <.001 |
| Co-authorship of scientific journal articles that use my patients' data | 91 (23.82) | 75 (30.99) | 10 (8.33) | <.001 |
| Only with my permission or the permission of my supervisor will researchers receive the clinical data of my patients (without identifying information) | 60 (15.70) | 45 (18.59) | 15 (12.50) | .142 |
| Mention my department/practice in scientific journal articles that use my patients' data | 37 (9.69) | 24 (9.92) | 13 (10.83) | .787 |
| Mention my name in the acknowledgements of scientific journal articles that use my patients' data | 13 (3.40) | 9 (3.80) | 3 (2.50) | .542 |

<sup>a</sup> Multiple Answer item; maximum 3 answers.
<sup>b</sup> Of all participants who answered this item (n=382), n= 369 indicated whether or not they consider themselves physician-scientists (n=242) or physicians (n=120).
<sup>c</sup> Chi-Squared Test (Pearson); values in italics are significant at .05 level of significance.

**Table 5:** Concerns about providing patients’ clinical data for secondary use and significance of group differences (n=382)

|  | all participants, n (%) | physician-scientists, n (%)<sup>b</sup> | Purely clinical physicians, n (%)<sup>b</sup> | *P value*<sup>c</sup> |
| --- | --- | --- | --- | --- |
| **Concerns about making patients' clinical data available<sup>a</sup>** |  |  |  |  |
| Misuse of data through unauthorized access to data records | 229 (59.94) | 127 (52.47) | 90 (75.00) | *<.001* |
| Insufficient data quality, which can cause studies to produce erroneous results | 215 (56.28) | 151 (62.39) | 50 (41.66) | *<.001* |
| Failure to adequately protect the privacy of my patients | 174 (45.54) | 87 (35.95) | 74 (61.66) | *<.001* |
| Other researchers conduct research using the clinical data from my practice/department before I do | 124 (32.46) | 104 (42.97) | 15 (12.50) | *<.001* |
| Novel liability issues/increased liability risk (e.g., research uncovers misdiagnosis in own practice/department) | 115 (30.10) | 62 (25.61) | 43 (35.83) | *.044* |
| New technological developments with new possibilities for re-identification carry the potential for harm to my patients | 89 (23.29) | 51 (21.07) | 32 (26.66) | *.233* |
| Discrimination against my patients on the basis of their clinical data, e.g. in the case of re-identification of individual patients or on the basis of belonging to a group with a certain disease or disposition | 74 (19.37) | 37 (15.28) | 32 (26.66) | *.009* |
| Performance comparisons with other practices/departments at hospitals conducted using clinical data from external agencies | 71 (18.58) | 47 (19.42) | 21 (17.50) | *.659* |
| Loss of trust in the doctor-patient relationship on the part of the patients | 61 (15.96) | 28 (11.57) | 25 (20.83) | *.019* |
| Future studies pursue research purposes that harm my patients | 31 (8.11) | 16 (6.61) | 11 (9.16) | *.384* |

<sup>a</sup> Multiple Answer item.
<sup>b</sup> Of all participants who answered this item (n=382), n= 369 indicated whether or not they consider themselves physician-scientists (n=242) or physicians (n=120).
<sup>c</sup> Chi-Squared Test (Pearson); values in italics are significant at .05 level of significance.

## Discussion

### Main Findings

Information on whether and under what preconditions physicians are willing to make their patients’ clinical data available for research is vital for practical and ethically sound implementation of secondary use. Here we report results of a survey among 397 physicians working in two university medical centres and in private practice on their general attitudes, concerns, in-principle willingness and conditions for enabling support of secondary use of clinical data for research purposes. To our knowledge, we present the first quantitative analysis of physicians’ conditions to support secondary use of clinical data for research purposes.

Firstly, we found a highly positive general attitude of physicians towards secondary use along with little fundamental reservations. Secondly, physicians showed widespread in-principle willingness to support secondary use; most important conditions for practical implementation were reliable protection of patients’ privacy as well as funding of additional person-hours for documentation and consenting patients. Third, group specific differences were prevalent: Physician-scientists were more likely to be concerned about data quality, required additional funding for research-adequate documentation and the right to first conduct research with their patients’ clinical data; in contrast, purely clinical physicians were more prone to be concerned about patient privacy and the physician-patient relationships, and to require, hence, reliable patient privacy as well as monetary compensation.

### High in-principle willingness for supporting secondary use

Despite methodological and thematic differences that limit comparability, we aim to embed our findings carefully within related studies. Among all 397 surveyed physicians, a very high proportion of physicians were of the view that secondary use is important (96%) and were in-principle willing to support secondary use (87%). This corresponds with a previous survey in a small sample in Canadian health researchers presenting the very strong general acceptance (96%) of using “citizens’ health data for research” (10). Our study further found a majority of physicians viewing the support of secondary use as a moral duty of peer physicians (68%). This view resonates with the results of a study we have previously conducted in which cancer patients attributed an obligation to their physicians to support secondary use (91%) (12). An ethical discussion of such a duty to support secondary use seems essential in order to resolve the tension of conflicting duties. Only few physicians expressed fundamental reservations about secondary use (8%). This finding helps to classify the importance of qualitative studies that emphasized widespread and severe concerns to the German context (4, 5, 11).

This study showed that making clinical data available for researchers who work for companies and conduct medical research was acceptable for the majority of physicians (40%). This aspect was previously considered problematic in qualitative studies among physicians (5, 8, 13). Acceptance rate of data use by company researchers in our study was considerably higher than in a small sample of GPs conducted in Canada being asked for use of electronic health record data for research by pharmaceutical industry (9%; n=46) (11). This discrepancy should be examined in a more nuanced way with a focus on types of data use and consideration of wordings such as ‘pharmaceutical industry’ which could carry negative connotations. Physicians were more willing to support secondary use in case of public-private-partnerships which is consistent with findings of a qualitative study with GPs conducted in the UK that indicate a higher level of trust in public-private partnerships than in companies (6).

### Most important conditions for practical implementation

#### Adjustment of workflows

About three-quarters of physicians were in principle willing to adjust their workflows to support secondary use (76%). Physicians were most interested in keeping expenditures in terms of time, personnel and money to a minimum. This is consistent with a Canadian qualitative study in GPs identifying uncompensated staff time as a major hurdle (5). Studies report that physicians being already increasingly dissatisfied with more and more time occupied for documenting in electronic health records for medical care – meaning even without documentation for research purposes - at the expense of time spent in contact with patients (14, 15). One quantitative study even suggests electronic health record use being related to burnout (16). Hence, physicians will likely be sensitive towards additional time and work that could further reduce contact time with their patients. While physicians in this study indicated that additional documentation time per patient visit of 5 minutes on average would be acceptable for them, they also specifically requested extra funding for personnel for research-adequate documentation of data (77%). Software solutions might be apt to reduce the burden of documentation, if they decrease time for documentation significantly (17), and cover a range of functional tasks (18). Healthcare personnel and hospital management should be directly involved in the development of new workflows and software at an early stage (4) to prevent disruption of complex workflows.

The majority of physicians agreed that non-medical staff should consent patients for secondary use (62%). We suppose that physicians clearly distinguish consent to interventional clinical trials from consent to *non-interventional* secondary research use. Hence, obtaining informed consent for secondary use by trained, non-medical staff by default is worth considering.

#### Provision of patient data

In contrast to physician-scientists, purely clinical physicians were less inclined to be in principle willing to make their patients’ clinical data available for research purposes (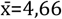 vs 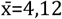, *P*<.001). Although comparability is limited due to differences in the study population this finding is in line with a survey among GPs from Canada with moderate acceptance rate of sharing patient data with university researchers (60%) (10).

Also, purely clinically oriented physicians were more likely to see patients as the owner of the data while overall the positions on the question whether data can be owned at all was quite divided. This reflects the controversial ethical debate about the concept of ownership with regard to health data where convincing arguments would rather support ownership in a sense of control and engagement than in the sense of property (19).

The most important condition on the part of physicians to make clinical data available for secondary use was the reliable protection of patient privacy (67%). This finding is consistent with existing literature indicating that physicians feel responsible for patient privacy and view themselves as data custodians (5, 20). Therefore, reliable data security and data protection should be implemented, the question of data ownership should be clarified, and physicians should be informed transparently about data protection measures of data initiatives (and also about data leaks if they occurred) so that trust in secondary use can be built.

Feedback systems that help physicians to improve internal quality and recognition of their efforts to contribute to a learning healthcare system might strengthen the link between care and research.

### Addressing physician-scientists’ research interests

We found that physician-scientists and purely clinical physicians differed systematically in terms of conditions they place on and concerns they have about secondary use. Physician-scientists’ research experiences and interests might be the reason for their significantly stronger concerns about insufficient data quality for research purposes. This finding is in line with a mixed-method study showing concerns about data quality among health researchers (21). To ensure physician-scientists’ trust in research-adequate data quality, implementing appropriate measures and resources for high quality documentation should receive high priority. Physician-scientists were also more likely than purely clinical physicians to demand additional funding for research-adequate documentation. According to literature, support for physician-scientists is needed to perform research activities alongside patient care (22).

Simultaneously, physicians should be made aware that their documentation practices have direct consequences for the scientific usability of clinical data and for the quality of research results.

Compared to purely clinical physicians, physician-scientists emphasised significantly more often the privilege to conduct research with patient data prior to other researchers. A systematic literature review demonstrated that health researchers want to exert some control over data they had collected (3). Hence, potential rights of – time-limited - exclusive use might facilitate the implementation of secondary use, but need to be weighed against the argument of maximizing utility of data generated in a publicly funded healthcare system. Additional measures might be co-authorship or other ways of recognition.

### Addressing purely clinical physicians’ interests

Purely clinical physicians were significantly more often concerned than physician-scientists about their patients loosing trust in the physician-patient relationship if data are made available for secondary use. Such concerns have not been reported so far. Purely clinical physicians were also more likely to be concerned about protection of their patients’ privacy and placed reliable privacy protection as the most important condition to support secondary use. They even reported to protect their patients’ data by documenting differently. According to the literature, trust in data users seems to be a vital facilitator for secondary use (5, 21, 23) as well as a trustworthy governance structure and oversight bodies, e.g. use and access committees (24) (13). To build trust in data infrastructure and governance of secondary use, ensuring and communicating patient privacy protection is a fundamental prerequisite, particularly for purely clinical physicians.

An important incentive for purely clinical physicians was monetary compensation of expenses for secondary use. Since almost all purely clinical physicians work in private practices, this finding might reflect their economic situation as mostly self-employed entrepreneurs. Since purely clinical physicians typically do not plan to use and directly benefit from the preparation and provision of data, fair compensation schemes seem imperative for this group (5).

## Limitations

The low response rate (5%) reflects difficulties to motivate physicians to participate in studies have been recognized (25) with reasons for non-participation such as survey fatigue and minimal time resources. Also, the subject of the present study does not directly address topics relevant to patient care which may have further reduced interest in participation. Given the low response rate, self-selection bias cannot be ruled out. The high proportion of physicians working at university hospitals might lead to a study population with increased thematic (research) interest, possibly overstating positive attitude towards secondary use. We used a self-developed questionnaire without validated measurement instruments, yet tested by cognitive interviews. The sample is not representative of the German medical profession, yet our results may provide indications of relevant needs and concerns of physicians in Germany.

We assumed that organizational background exerts relevant influence on physicians’ attitudes toward secondary use as physicians working in private practice potentially feel more in charge for data protection standards of their practice than physicians working in a hospital. The distinction between physicians working in hospitals and in private practice needs further assessment in a larger dataset to inform implementation of secondary use in different organisational settings.

## Conclusion

This first quantitative study on the perspective on secondary health data use of physicians in an research prone environment compared to those in private practice should inform further studies and the setup of infrastructures for secondary use of clinical data in Germany and possibly beyond. We found high in-principle willingness of physicians to support secondary use and low general concerns. High in-principle willingness and little concerns indicate the importance of considering physicians’ demands and conditions in order to foster secondary use: most important conditions were protection of patient privacy and manageable expenses in terms of time, personnel and money. If extra expenses occur, the provision of funding to compensate for them is expected such as medical documentation specialists, non-medical staff obtaining consent, and user-centred documentation software - in order to not further reduce contact time with patients. Adaptation of workflows for research-adequate documentation and consenting patients should be pilot-tested in participatory (research) formats in order to prevent disruption of complex clinical processes.

Our results demonstrated group-specific differences. *Physician-scientists’* answers mirrored the rationales of the scientific system with concerns about research-adequate data quality, requesting incentives such as the privilege of first conducting research and funding for research-adequate documentation. Building trust in data repositories and its users seems essential for physician-scientists’ support and readiness to conduct research with clinical data themselves. *Purely clinical physicians* were concerned about patients’ privacy and about a possibly worsening physician-patient relationship. Their most important condition for support of secondary use was the protection of patient privacy but also monetary compensation which can be attributed to the often self-employed work in private practices performed by this group. Besides establishing monetary compensation schemes, for purely clinical physicians, relevant conditions to support secondary use include ensuring and communicating patient privacy protection accompanied by a trustworthy data governance structure that enables transparent data use.

## Data Availability

All data files (dataset, information about data cleansing, and questionnaire) are available from the heiDATA database (accession number UNF:6:rCJxTnlT0ZCX6/Tn/Zv78g==doi: https://heidata.uni-heidelberg.de/dataset.xhtml?persistentId=10.11588/data/5JCEVW).

## Ethics approval and consent to participate

Informed consent was obtained from individuals who participated in the study pretests and the written survey. The datasets generated and analyzed in the current study are available online (https://doi.org/10.11588/data/5JCEVW).

This manuscript was developed within the framework of the project “Learning from Clinical Data (LinCDat)” funded by the Deutsche Forschungsgemeinschaft (DFG, German Research Foundation) – 406103282. The project funding was awarded to ECW. The funders had neither involvement in study design; in the collection, analysis and interpretation of data; in the writing of the report, nor in the decision to submit the article for publication. There was no additional external funding received for this study.

All authors contributed to the study conception and design. Material preparation, data collection and analysis were performed by AK and KM. ECW and KM supervised the work and supported data interpretation. The first draft of the manuscript was written by AK and iteratively reviewed by ECW. All authors commented on previous versions of the manuscript. All authors read and approved the final manuscript.

We would like to thank the Cancer Registry of Baden-Württemberg for support of data collection as well as Sein Schmidt and Verena Benz of the Charité-BIH Clinical Study Center. We also thank our project partners Kai Cornelius, Markus Spitz (University of Heidelberg, Germany) and Adrian Thorogood (University of Luxemburg) for conceptual counselling. We are also thankful to Kai Cornelius for establishing contact with the Charité-BIH Clinical Study Center We acknowledge Hellen Kachler and Niamh Sulzbach (both: National Center for Tumor Diseases, Heidelberg University Hospital, Germany) for their assistance in researching the literature, data entry, and linguistic correction of the manuscript.

